# Clinical Performance Evaluation of a SARS-CoV-2 Rapid Antibody Test for Determining Past Exposure to SARS-CoV-2

**DOI:** 10.1101/2020.09.01.20180687

**Authors:** Peter Findeisen, Hugo Stiegler, Eloisa Lopez-Calle, Tanja Schneider, Eva Urlaub, Johannes Hayer, Claudia Zemmrich

**Affiliations:** MVZ Labor Dr. Limbach & Kollegen GbR, Heidelberg, Germany; MVZLM - Medizinisches Versorgungszentrum für Labormedizin und Mikrobiologie Ruhr GmbH, 45138 Essen, Germany; Roche Diagnostics GmbH, Mannheim, Germany; Roche Diagnostics GmbH, Penzberg, Germany; Institute for Pharmacology and Preventive Medicine, Menzelstrasse 21, 15831 Mahlow, Germany; Roche Diagnostics International Ltd. Rotkreuz, Switzerland

**Keywords:** SARS-CoV-2, rapid antibody test, past exposure

## Abstract

The true prevalence and population seropositivity of SARS-CoV-2 infection remains unknown, due to the number of asymptomatic infections and limited access to high-performance antibody tests. To control the COVID-19 pandemic it is crucial to understand the true seroprevalence, but not every region has access to extensive centralized PCR and serology testing. Currently available rapid antibody tests lack the accuracy needed for recommendation by health authorities. To fill this gap, we analyzed and validated the clinical performance of a new point-of-care SARS-CoV-2 Rapid Antibody Assay, a chromatographic immunoassay for qualitative detection of IgM/IgG antibodies for use in near-patient settings. Analysis was performed using 42 Anti-SARS-Cov-2 positive (CoV+) and 92 Anti-SARS-Covid-2 negative (CoV-) leftover samples from before December 2019, using the Elecsys® Anti-SARS-CoV-2 as the reference assay. Analytical specificity was tested using leftover samples from individuals with symptoms of common cold collected before December 2019. The SARS-CoV-2 Rapid Antibody Test was 100.0% (95% CI 91.59–100.00) sensitive and 96.74% (95% CI 90.77–99.32) specific with an assay failure rate of 0.00%. No cross-reactivity was observed against the common cold panel. Method comparison was additionally conducted by two external laboratories, using 100 CoV+/275 CoV-samples, also comparing whole blood versus plasma matrix. The comparison demonstrated for plasma 96.00% positive/96.36% negative percent agreement with the Elecsys Anti-SARS-CoV-2 and overall 99.20% percent agreement between whole blood and EDTA plasma. The SARS-CoV-2 Rapid Antibody Test demonstrated similar clinical performance to the manufacturer's data and to a centralized automated immunoassay, with no cross-reactivity to common cold panels.

## Introduction

The global COVID-19 pandemic created an urgent unmet clinical need to investigate reliable diagnostic tools for patients, as well as understand the extent of exposure and spread of infection among wider populations (1-3). Acute diagnosis of the COVID-19 infection is based on identification of viral RNA via PCR from swab samples, which is detectable from symptom onset for approximately four weeks (2, 4). As is known from localized testing during outbreaks, many people who are infected with the virus do not present with any clinical symptoms - current estimates suggest around 30% of seropositive individuals are asymptomatic (5-7). Those individuals carry the virus and potentially spread it to others, who may react with a severe COVID-19 disease (8). No region in the world can perform PCR testing of every patient with common cold symptoms or who has had contact with a suspected COVID-19 patient. In addition to clinical testing of individuals with suspected COVID-19 for direct virus detection, surveillance strategies need to combine several diagnostic techniques to monitor disease kinetics in wider populations (2, 9). To control the pandemic, it seems crucial to investigate who has already had an infection and has developed antibodies as an immune response, and vice versa, who is still vulnerable to an infection (10-14).

Antibody tests are not intended to diagnose an acute COVID-19 infection, more specific diagnostic methods should be performed to obtain this (2). Ongoing research on the level and duration of immunity of seropositive persons will add further value to the clinical and epidemiological interpretation of positive antibody testing results. Preliminary data suggest that high affinity antibody tests show good correlation with neutralizing activity (15).

Based on current evidence, immunoglobulin M (IgM) antibodies are detectable within 5 days after symptom onset and immunoglobulin G (IgG) antibodies within 5–7 days (4, 16-23). Depending on the applied method, seroconversion is observed after a median of 10–13 days after symptom onset for IgM and 12–14 days for IgG, and maximum for both is reached after 2 weeks (4, 16, 19-26). Individual levels and kinetics of both IgM and IgG are highly variable, which is why simultaneous detection of both is recommended.

Currently available rapid antibody tests require improved accuracy before being recommended by competent authorities and used by healthcare professionals (HCPs) in the wider population (2, 3). The US Food and Drug Administration (FDA) released technical requirements for antibody tests on April 4, 2020, that include a specificity of ≥ 95% and cross-reactivity testing for common cold and other coronaviruses (27). There are different high-throughput Anti-SARS-CoV-2 antibody tests available; the assay selected as the reference for our test (Elecsys Anti-SARS-CoV-2 Immunoassay) is based on electrochemiluminescence (ECLIA), using a doubleantigen sandwich test principle and a recombinant protein representing the antigen for the determination of antibodies to SARS-CoV-2, namely the nucleocapsid protein (N) (28). It provides a qualitative result with a sensitivity of 100.0% (95% CI 88.10–100.0) at > 14 days after PCR confirmation and a specificity of 99.81% (95% CI 99.65–99.91) (28).

Rapid tests, also called point-of-care (PoC) tests, combine immunoassay and chromatography for a qualitative detection of antibodies. The selected test *(Anti-SARS-CoV-2 Rapid Antibody Test [SD Biosensor, Chungcheongbuk-do, Republic of Korea])* is a CE marked lateral flow assay displaying a visual ‘yes/no’ answer for the selective detection of specific IgG and/or IgM antibodies to SARS-CoV-2, with two separate colored bands for IgG and IgM (29). The manufacturer states the specificity is 98.65% and sensitivity beyond 14 days after symptom 4 onset of 99.03%, tested on 103 PCR-confirmed CoV+ and 222 CoV-samples (29). The samples had also been tested for early sensitivity between 7 and 14 days after symptom onset with a result of 92.59%. SD Biosensor has performed several cross-reactivity studies involving numerous specimens, including influenza A and B.

In this study series, we further validated and extended the manufacturers' clinical performance and cross-reactivity data and externally performed a matrix and method comparison to gain additional data on the overall assay performance.

## Materials and Methods

### The assay

The assay ***(Figure 1)*** needs 10 μl human serum or plasma, or 20 μl whole venous or capillary blood sample to be filled into the preformed well of the test device. Three pre-coated lines mark the “C” control line, and the “G” and “M” test lines for IgG and IgM. Monoclonal chicken IgY antibody is coated on the “C” region, and monoclonal anti-human IgG antibody and monoclonal anti-human IgM antibody are coated on the “G” and “M” test line region. During the test, SARS-CoV-2-specific antibodies in the sample interact with recombinant SARS-CoV-2 protein (nucleocapsid and spike protein) conjugated with colloidal gold particles forming antibody-antigen gold particle complexes. This complex migrates on the membrane via capillary action until the “M” and “G” test line, where it will be captured by the monoclonal anti-human IgG antibody or monoclonal anti-human IgM antibody. A violet test line would be visible in the result window if SARS-CoV-2-specific antibodies are present in the sample. The intensity of the colored test line varies depending upon the amount of SARS-CoV-2 antibodies present in the sample. Even if the color is faint, the test result should be interpreted as a positive result. The control line is used as procedural control and should always appear if the test procedure is performed properly and the test reagents are working.

**Figure 1.**
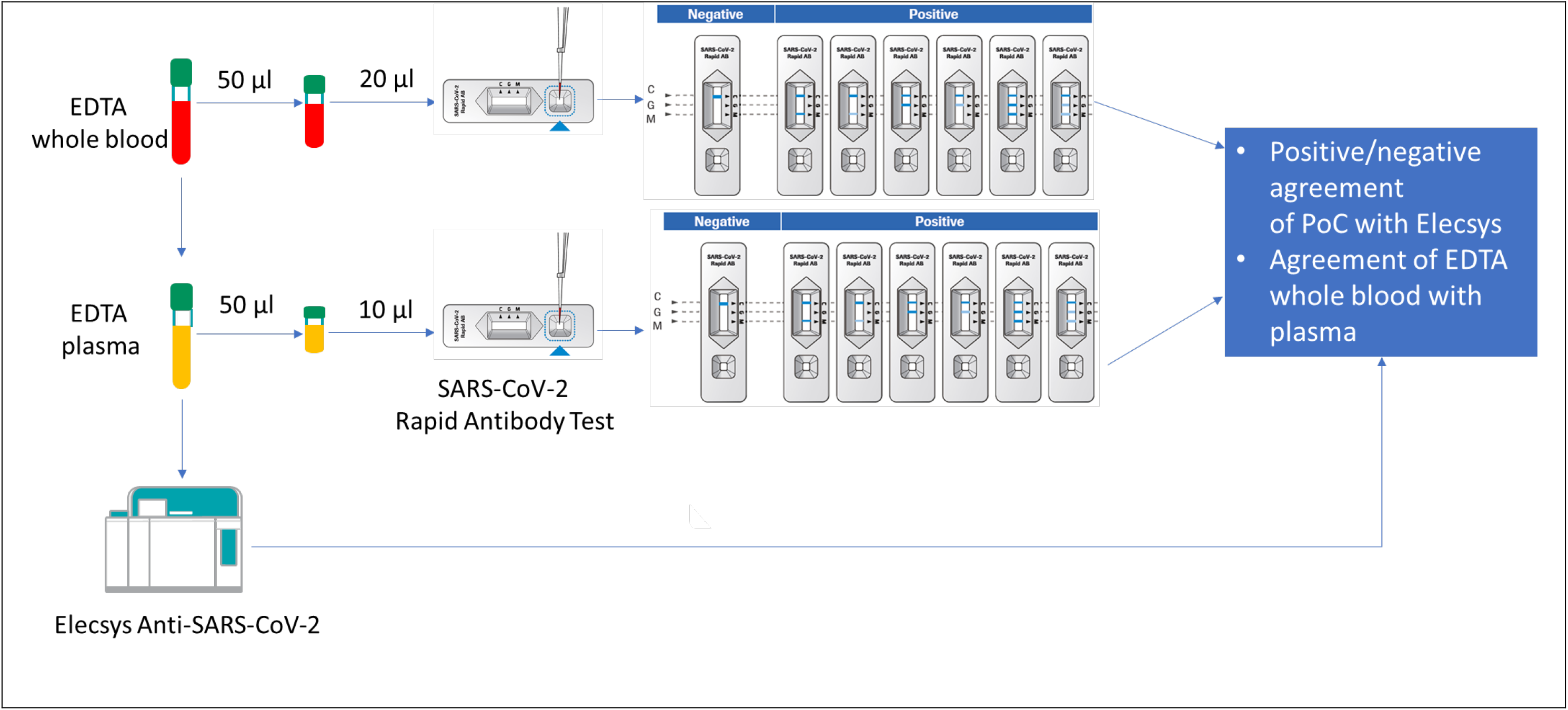
External method and matrix comparison - workflow.

### Study design

A performance analysis was conducted at Roche Diagnostics (Penzberg, Mannheim, Germany) using 42 Elecsys Anti-SARS-CoV-2 confirmed CoV+ and 92 leftover samples from healthy donors, collected before December 2019 and additionally confirmed by the Elecsys Anti-SARS-CoV-2 CoV-(56 lithium heparin plasma, 36 EDTA plasma). Cross-reactivity testing was conducted with 18 samples from individuals expressing signs and symptoms of common cold (i.e. sore throat, cough, fever) collected before December 2019. Additional matrix equivalence and read-out time analysis captured 159 Elecsys referenced CoV-samples, consisting of 55 heparin plasma, 55 EDTA plasma and 49 serum samples.

Additionally, an independent comparison was performed with matched EDTA plasma and whole blood samples from a total of 375 anonymized leftover samples at two external testing sites. In total, 100 samples tested positive for SARS-CoV-2 antibodies by the Elecsys anti-SARS-CoV2 immunoassay and 275 negative samples (25 CoV+/75 negative-subjects at MVZLM Ruhr GmbH, Essen and 75 CoV+/200 negative subjects at MVZ Labor Dr. Limbach & Kollegen GbR Heidelberg). No information on PCR result or time of sample collection related to symptom onset was available. All samples were analyzed with the Anti-SARS-CoV-2 Rapid Antibody Assay and results were compared directly with the EDTA plasma sample from the Elecsys Anti-SARS-CoV-2 Assay (see Figure 1 for workflow).

All investigations were performed according in a single determination and according to the manufacturer's instructions for use.

### Statistical analyses

For sensitivity and specificity, point estimates and 95% CI values were calculated. To determine positive and negative percent agreement (PPA and NPA), the Elecsys Anti-SARS-CoV-2 result was used as comparator: »Non-reactive« (cut off index [COI] < 1.0) was a negative result and »Reactive« (COI ≥ 1.0) a positive result. The rapid test was considered positive in case either IgG or IgM showed a colored line, even if the line was faint. The Clopper-Pearson exact method was used for the calculation of the two-sided 95% confidence intervals (CIs).

### Ethics and conformity declaration

The study was conducted in accordance with applicable regulations, including relevant European Union directives and regulations, and the principles of the Declaration of Helsinki. All samples were anonymized leftover specimens. For the samples tested at Roche Diagnostics, a statement was obtained from the Ethics Committee of the Landesärztekammer Bayern confirming that there are no objections against the transfer and the coherent use of the anonymized leftover samples. For the samples tested in MVZLM Ruhr GmbH (Essen) and at MVZ Labor Dr. Limbach & Kollegen GbR (Heidelberg) no ethics committee vote is required in accordance with MPG (Medizinproduktegesetz Deutschland).

### Data availability

Qualified researchers may request access to individual patient level data through the clinical study data request platform (https://vivli.org/). Further details on Roche's criteria for eligible studies are available here: https://vivli.org/members/ourmembers/. For further details on Roche's Global Policy on the Sharing of Clinical Information and how to request access to related clinical study documents, see here: https://www.roche.com/research_and_development/who_we_are_how_we_work/clinical_trials/our_commitment_to_data_sharing.htm.

## Results

### Clinical performance evaluation

The assay presented a qualitative visual test result without the need for a read-out instrument like other rapid antibody devices on the market. Some lines were quite faint, and according to manufacturer's instruction they have been interpreted as a positive result. Handling was easy with one sample-transfer step and three drops of buffer dropped into the well after the blood sample; the results read at 10–15 minutes. Except quality control material, lancets and a transferring pipette for 10 |al, everything needed to conduct the test was included into the test package. All test cassettes were correctly assembled, opening the foil pouch was easy without danger of destroying the desiccant and no membranes were scratched.

A total of 42 left-over samples with prior Elecsys-confirmed positive SARS-CoV-2 antibody detection were included in the sensitivity analysis. The overall sensitivity was 100.0% (95 % CI 91.59–100.0) (Table 1). A total of 92 samples from healthy donors before December 2019 presented an overall specificity of 96.74% (95% CI 90.77–99.32) with no difference between EDTA and lithium-heparin plasma: 97.22% (95% CI: 85.47–99.93)/96.43% (95% CI 87.69–99.56).

**Table 1.**
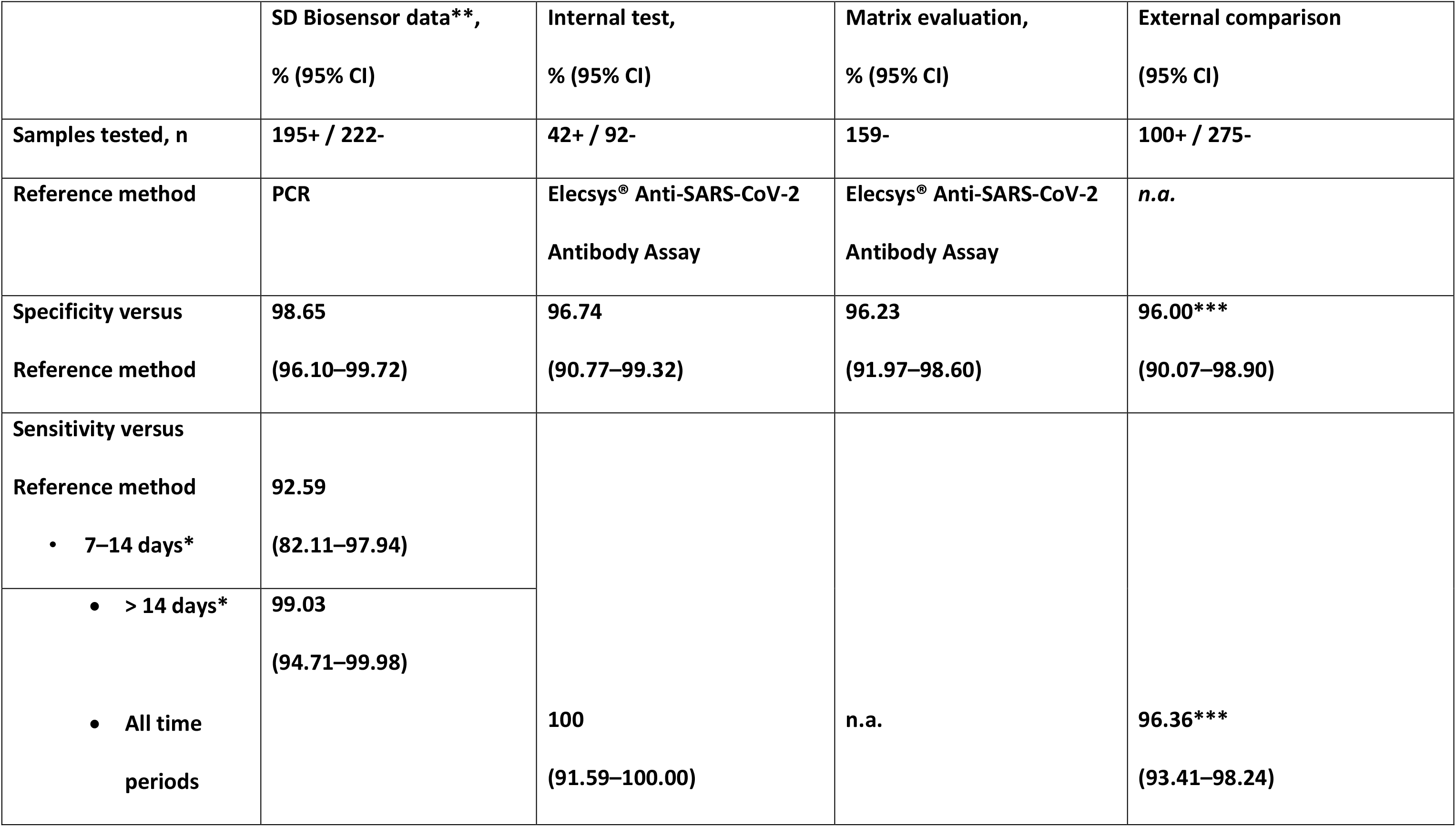
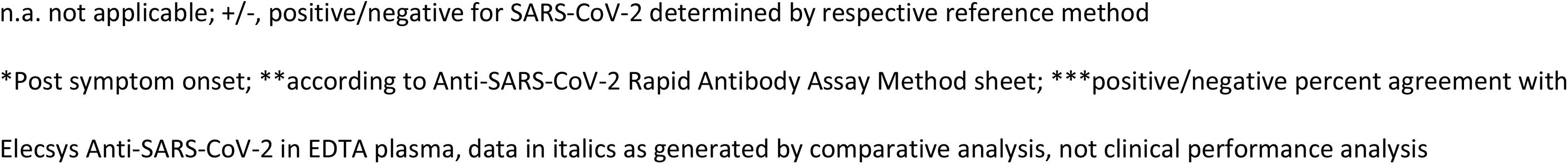
Performance data of the manufacturer, internal clinical performance test, matrix equivalence test and external method comparison.

An additional matrix evaluation and read-out time analysis was performed with 159 SARS-CoV-2 negative samples to confirm equal results throughout the pre-defined read-out time window. At read out times ≤10 minutes specificity was slightly higher versus at 15 minutes (Table 2), however detectable signals were ‘weaker’ or less well defined at ≤10 minutes compared with 15 minutes read out time. No relevant performance differences were noticed between serum, heparin and EDTA plasma (Table 2).

**Table 2.** Matrix evaluation and read-out time analysis.

|  |  | Read out time |  |  |  |
| --- | --- | --- | --- | --- | --- |
|  |  | 8 mins | 10 mins | 12 mins | 15 mins |
| **Total** | **Number of samples test (negative)** | 159 | 159 | 159 | 159 |
|  | **False positive** | 4 | 5 | 6 | 6 |
|  | **Specificity % (95% CI)** | 97.48 (93.68–99.31) | 96.86 (92.81–98.97) | 96.23 (91.97–98.60) | 96.23 (91.97–98.60) |
| **Serum** | **Number of samples test (negative)** | 49 | 49 | 49 | 49 |
|  | **False positive** | 1 | 2 | 2 | 2 |
|  | **Specificity % (95% CI)** | 97.96 (89.15–99.95) | 95.92 (86.02–99.50) | 95.92 (86.02–99.50) | 95.92 (86.02–99.50) |
| **Heparin**<br>**plasma** | **Number of samples test (negative)** | 55 | 55 | 55 | 55 |
|  | **False positive** | 1 | 1 | 2 | 2 |
|  | **Specificity % (95% CI)** | 98.18 (90.28–99.95) | 98.18 (90.28–99.95) | 96.36 (87.47–99.56) | 96.36 (87.47–99.56) |
| **EDTA**<br>**plasma** | **Number of samples test (negative)** | 55 | 55 | 55 | 55 |
|  | **False positive** | 2 | 2 | 2 | 2 |
|  | **Specificity % (95% CI)** | 96.36 (87.47–99.56) | 96.36 (87.47–99.56) | 96.36 (87.47–99.56) | 96.36 (87.47–99.56) |

Cross-reactivity was tested at 18 samples from a common cold panel, collected before December 2019 without further information on the exact pathogens of the specimens. For all 18 measurements, a colored control line was obtained, indicating that the test worked properly. On the test lines for IgG and IgM no color appeared, thus leading to a negative result and resulting in a specificity of 100.0% (95 % CI 81.47–100.0%). The results demonstrated that the test is not reacting with antibodies directed against related pathogens from common cold infections.

### External method and matrix comparison

Total results for both testing methods (plasma/whole blood samples) were 96.00% (95 % CI 90.07–98.90) / 94.00% (95% CI 87.40–97.77) for positive percent agreement rate and 96.36% (95% CI 93.41–98.24) / 96.00 % (95% CI 92.96–97.99) for negative percent agreement rate. The sample numbers per matrix and per immunoglobulin class for the samples confirmed by the Elecsys test and the numbers differing from the Elecsys result are summarized in Table 3. The overall percent agreement rate between whole blood and EDTA plasma was 99.20% (95% CI 97.68–99.83), the positive/negative percent agreement rates for whole blood versus EDTA plasma were 98.11 % (95% CI 93.35–99.77) / 99.63 (95% CI 97.95–99.99).

**Table 3.** External comparison to Elecsys Anti-SARS-CoV-2 per matrix and immunoglobulin.

|  | Number of results (Whole blood) |  |  |  | Number of results (EDTA Plasma) |  |  |  |
| --- | --- | --- | --- | --- | --- | --- | --- | --- |
|  | IgM | IgG | IgM+IgG | Total | IgM | IgG | IgM+IgG | Total |
| **SARS-CoV-2 positive (confirmed by Elecsys)** | 1 | 57 | 36 | 94 | 0 | 54 | 42 | 96 |
| **SARS-CoV-2 negative (confirmed by Elecsys)** | 0 | 0 | 0 | 264 | 0 | 0 | 0 | 265 |
| **False positive results (compared to Elecsys)** | 4 | 5 | 2 | 11 | 3 | 5 | 2 | 10 |
| **False negative results (compared to Elecsys)** | 0 | 0 | 0 | 6 | 0 | 0 | 0 | 4 |
| **OPA (95%CI)** | 95.47% (92.84–97.34) |  |  |  | 96.27% (93.82–97.94) |  |  |  |
| **PPA (95% CI)** | 94.00% (87.40–97.77) |  |  |  | 96.00% (90.07–98.90) |  |  |  |
| **NPA (95% CI)** | 96.00% (92.96–97.99) |  |  |  | 96.36% (93.41–98.24) |  |  |  |

OPA, overall percentage agreement; PPA, positive percentage agreement; NPA, negative
percentage agreement

A total of 10 EDTA samples and 11 whole blood samples were detected antibody positive by the Anti-SARS-CoV-2 Rapid Antibody Assay but antibody negative by the Elecsys assay, see Table 4a for the respectively detected immunoglobulin classes and the respective signal intensity on the 11 rapid tests. 96 plasma and 94 whole blood samples out of 100 Elecsys-positive samples were detected positive by the rapid test, see Table 4b for the respective immunoglobulin classes and signal intensity of the six rapid test results. The matrix comparison for the Anti-SARS-CoV-2 Rapid Antibody Assay found of 106 plasma samples with an antibody positive test result, included two samples which had negative tests results with whole blood. One out of 269 negative test results on the rapid test with plasma displayed a positive result with whole blood.

**Table 4a:** Method comparison: Discrepant positive samples - detected signal intensity and immunoglobulin class.

| Sample number | Rapid AB results – whole blood |  | Rapid AB results – EDTA plasma |  | Elecsys results (EDTA plasma) |  |
| --- | --- | --- | --- | --- | --- | --- |
|  | IgG | IgM | IgG | IgM | Result | COI |
| 1 | - | X (weak) | - | X (weak) | Non-reactive | 0.089 |
| 2 | - | X (weak) | - | X (weak) | Non-reactive | 0.119 |
| 3 | - | X (weak) | - | X (weak) | Non-reactive | 0.089 |
| 4 | X (weak) | - | X (weak) | - | Non-reactive | 0.079 |
| 5 | - | X (weak) | - | - | Non-reactive | 0.069 |
| 6 | X (weak) | X (weak) | X (weak) | X (weak) | Non-reactive | 0.229 |
| 7 | X | - | X | - | Non-reactive | 0.146 |
| 8 | X (weak) | - | X (weak) | - | Non-reactive | 0.221 |
| 9 | X (weak) | - | X (weak) | - | Non-reactive | 0.269 |
| 10 | X | X | X | X | Non-reactive | 0.189 |
| 11 | X | - | X | - | Non-reactive | 0.378 |

AB, antibody

**Table 4b.** Method comparison: Discrepant negative samples - detected signal intensity and immunoglobulin class.

| Sample<br>number | Rapid AB results – whole blood |  | Rapid AB results – EDTA plasma |  | Elecsys results (EDTA plasma) |  |
| --- | --- | --- | --- | --- | --- | --- |
|  | IgG | IgM | IgG | IgM | Result | COI |
| 1 | - | - | - | - | Reactive | 1.21 |
| 2 | - | - | - | - | Reactive | 1.99 |
| 3 | - | - | X (weak) | - | Reactive | 39.54 |
| 4 | - | - | - | - | Reactive | 2.32 |
| 5 | - | - | - | - | Reactive | 2.34 |
| 6 | - | - | X (weak) | - | Reactive | 16.7 |

AB, antibody

## Discussion

The COVID-19 pandemic has created an urgent need for antibody testing of large populations to determine seroprevalence and potential immunity, which will be of more importance if more conclusive scientific data on correlation between these factors become available (2, 3, 26, 30, 31). The anticipated need for high testing capacities in PoC settings outside of large hospitals, calls for the development and validation of high-performance rapid antibody tests as reliable diagnostic instruments, in addition to the centralized antibody tests available. The evaluated Anti-SARS-CoV-2 Rapid Antibody Test complies with the acceptance criteria, defined by the FDA EUA on April 4, 2020, for SARS-CoV-2 antibody test developments (27). Our internal clinical performance evaluation confirmed the manufacturers' reported clinical sensitivity of the test with an uncharacterized cohort of antibody positive samples from a population with unknown time from symptom onset, which may reflect the typical clinical scenario for the test indication. The external comparison confirmed high agreement of both the Elecsys assay and the Anti-SARS-CoV-2 Rapid Antibody Test, also with no confirmed PCR result and time of sampling from symptom onset unknown. For both the internal and external evaluation, samples could be mixed, with some collected early after symptom onset. Both tests claim less than 100 % sensitivity <14 days after symptom onset (28, 29). The Elecsys Anti-SARS-CoV-2 Antibody Test measures high-affinity IgG/IgM antibodies directed at the nucleocapsid antigen, with an excellent sensitivity of 100.0% (95% CI 88.1–100.0) at ≥ 14 days after PCR confirmation (28). The rapid test has been developed detecting both nucleocapsid- and spike protein-associated antibodies to increase test accuracy compared with centralized serological assays, which typically perform better than rapid tests. (29). A limitation of this study is that samples were not differentiated according to time of collection after symptom onset, as such some of the false positive samples might actually be true positives, but were not detected by the Elecsys assay and most likely indicate an early antibody response with low affinity antibodies. For SARS-CoV-2 tests high specificity is a priority, particularly in low prevalence settings (30), as such it would be interesting to further evaluate the interpretation of the low positive samples at the rapid test, particularly for samples that were IgM positive only, to determine if these reflect non-specific binding, or true early detection. Further assessment with serial samples taken from COVID-19 patients from early infection phase onwards will help to better understand this.

The investigated Anti-SARS-CoV-2 Rapid Antibody Test provided readily evaluable results, regardless of sample type (whole blood or EDTA plasma) and independent of any read-out time between 10 and 15 minutes, as stated in the instructions for use. Those positive practical aspects could enable potential use outside medical environments. The assay is currently intended for professional use in laboratory and PoC environments as an aid in identifying individuals with an adaptive immune response to SARS-CoV-2, indicating prior infection.

In these studies, the SARS-CoV-2 Rapid Antibody Test demonstrates excellent clinical performance without cross-reactivity to common cold samples and results comparable to data of an automated high-performance immunoassay (Elecsys). Our data confirm and extend the manufacturers' performance data and add further details on matrix and method comparisons. Available data support its use as a reliable diagnostic instrument for SARS-CoV-2 IgM and IgG antibody detection in near-patient settings with potential extended usability outside medical environments.

## Acknowledgements

The authors would like to acknowledge the contributions of Rabea Held, Regina Draude, Marcus Pollok (Roche Diagnostics GmbH, Mannheim, Germany) for testing, data acquisition in the laboratory and preliminary data analysis. In addition, thanks to Imola Szebenyi, Oscar Gutierrez-Sanz, Miriam Hübner, Ludwig Pollich, Nicolai Bluthardt, Andreas Aristow (Roche Diagnostics GmbH, Penzberg, Germany) for the measurements of the clinical and crossreactivity samples as part of the study conducted at Roche Diagnostics. Thanks to John Burden (Roche Diagnostics International Ltd. Rotkreuz, Switzerland), Sven Groesgen (Roche Diagnostics GmbH, Mannheim, Germany) and Christine Jung (Roche Diagnostics GmbH, Penzberg, Germany) for planning, set-up and analysis of the external evaluation. The authors would also like to acknowledge Matthias Metz and Daria Glukhova for conducting the measurements with the Elecsys SARS-CoV-2 method as part of the study at Roche Diagnostics.

